# Genetic nurture and direct genetic transmission effects on body mass index across age

**DOI:** 10.64898/2026.08.31.26361795

**Authors:** Victória Trindade Pons, Nathan Gillespie, Roelof A.J. Smit, Joshua D. Arias, Xianyong Yin, Sonja I. Berndt, GIANT consortium, Albertine J. Oldehinkel, Hanna M. van Loo

## Abstract

Obesity is a growing public health challenge, with body mass index (BMI) influenced by both genetic and environmental factors. While the role of direct genetic transmission is well established, evidence for genetic nurture effects, in which parental genotypes impact offspring through the environment, has remained mixed. This study investigates direct genetic transmission and genetic nurture effects on BMI across ages, using parent-offspring trios and pairs from the Dutch Lifelines cohort study (N = 18,897 offspring, aged 8–67 years). We leveraged the latest multi-ancestry BMI polygenic score (PGS) to construct transmitted (PGS-T) and non-transmitted (PGS-NT) polygenic scores, where PGS-NT consists of parental alleles not passed on to offspring and serves as a proxy for genetic nurture. Linear mixed models showed a large effect of PGS-T on offspring BMI (β = 0.416, p < 0.001), corresponding to a 1.85 kg/m² increase per SD increase in PGS-T. PGS-NT had a small but significant effect (β = 0.026, p = 0.013), consistent with a genetic nurture effect accounting for approximately 6.6% of the effect of direct transmission. Parent-of-origin analyses showed that maternal PGS-NT effects were larger than paternal effects. PGS-T interactions with age indicated that direct transmission effects increased in childhood and stabilized in adulthood, while PGS-NT effects remained stable across age. Our findings suggest that direct genetic transmission is the dominant influence on BMI, while results are consistent with small genetic nurture effects that are driven by the maternal side.

## Introduction

Obesity rates are rising globally, posing significant public health challenges^1^. Body mass index (BMI), a common indicator of adiposity, is influenced by both genetic and environmental factors. Parents and offspring show moderate BMI correlations, with similarities persisting into adulthood^2^. This familial resemblance partly reflects the direct transmission of BMI-associated genes, as BMI is a moderately heritable trait, with estimates ranging from 40–90% in family studies^3^. However, parental genes for BMI may also shape the home environment through behaviors and habits that influence offspring BMI, such as dietary habits and physical activity routines^4,5^. This phenomenon, known as genetic nurture, where parental genotypes impact offspring phenotypes through the rearing environment rather than direct genetic inheritance^6,7^, provides a mechanism linking genetic predisposition and environmental influences on BMI. Disentangling the relative contribution of direct transmission and genetic nurture effects can help explain the familial clustering of obesity.

Research has shown that the heritability of BMI varies across development, with genetic influences increasing during childhood and stabilizing or decreasing from young adulthood into older age^8^. While the impact of direct transmission is established, evidence for genetic nurture effects on BMI has remained mixed. Early studies did not detect genetic nurture effects on BMI, overweight, or obesity^9,10^, but these were limited by small sample sizes and underpowered PGSs. More recently, one study reported significant genetic nurture effects on BMI that did not differ by age or parent^11^, while two others suggested these effects to be age-dependent and have a stronger maternal contribution^12,13^. Addressing these inconsistent patterns requires larger datasets with stronger genetic predictors, which can test whether genetic nurture effects on BMI vary by age, differ between mothers and fathers, and, critically, persist beyond adolescence.

Recent advances in genomic research now enable us to address these inconsistent findings about genetic nurture effects on BMI more definitively, as genome-wide association studies (GWAS) have now reached sample sizes over 5 million individuals^14^. This advancement enabled polygenic scores (PGS) to capture a significant proportion of BMI variation, with the latest multi-ancestry BMI-PGS explaining up to 17.6% of the variance in BMI in the UK Biobank. In this study, we leverage this PGS to construct transmitted (PGS-T) and non-transmitted (PGS-NT) parental haplotype-based scores^15^ to examine direct transmission and genetic nurture effects on BMI. By comparing the effects of PGS-T, which captures both direct genetic influences and potential genetic nurture effects, with PGS-NT, which serves as a proxy for the genetic nurture component, we can quantify the relative contribution of each mechanism to BMI development across the lifespan.

We analyzed BMI in offspring, initially aged 8 to 67 years, from parent-offspring trios and pairs from a large general population cohort in the Netherlands, with repeated BMI measurements over a decade. We expected that both PGS-T and PGS-NT would be positively associated with offspring BMI, reflecting direct genetic effects and genetic nurture effects, respectively. We examined whether these effects differed by parent-of-origin by separating the PGS by maternal and paternal haplotypes, and whether the associations were modified by age and sex.

## Results

### Sample characteristics

Lifelines included 18,897 offspring, of whom 3,227 were part of genotyped trios, 5,916 of father-offspring pairs, and 9,754 of mother-offspring pairs. After applying exclusion criteria, 42,184 BMI assessments were available, an average of 2.24 measurements per individual. Information on offspring BMI distribution is available in the Supplementary Information.

The mean BMI for the overall sample was 24.43, with offspring from parent-offspring trios showing a slightly lower mean BMI (23.92) than those from parent-offspring pairs (24.53). Offspring in trios were also younger at first assessment (mean age = 26.14) compared to those in pairs (mean age = 28.82). The proportion of females was 59.5% in the full sample, with a slightly higher proportion in pairs (60.1%) than in trios (56.7%). There were no significant differences in the mean transmitted PGS between trios and pairs (p=0.196). Detailed characteristics are presented in **Table 1**.

**Table 1.**
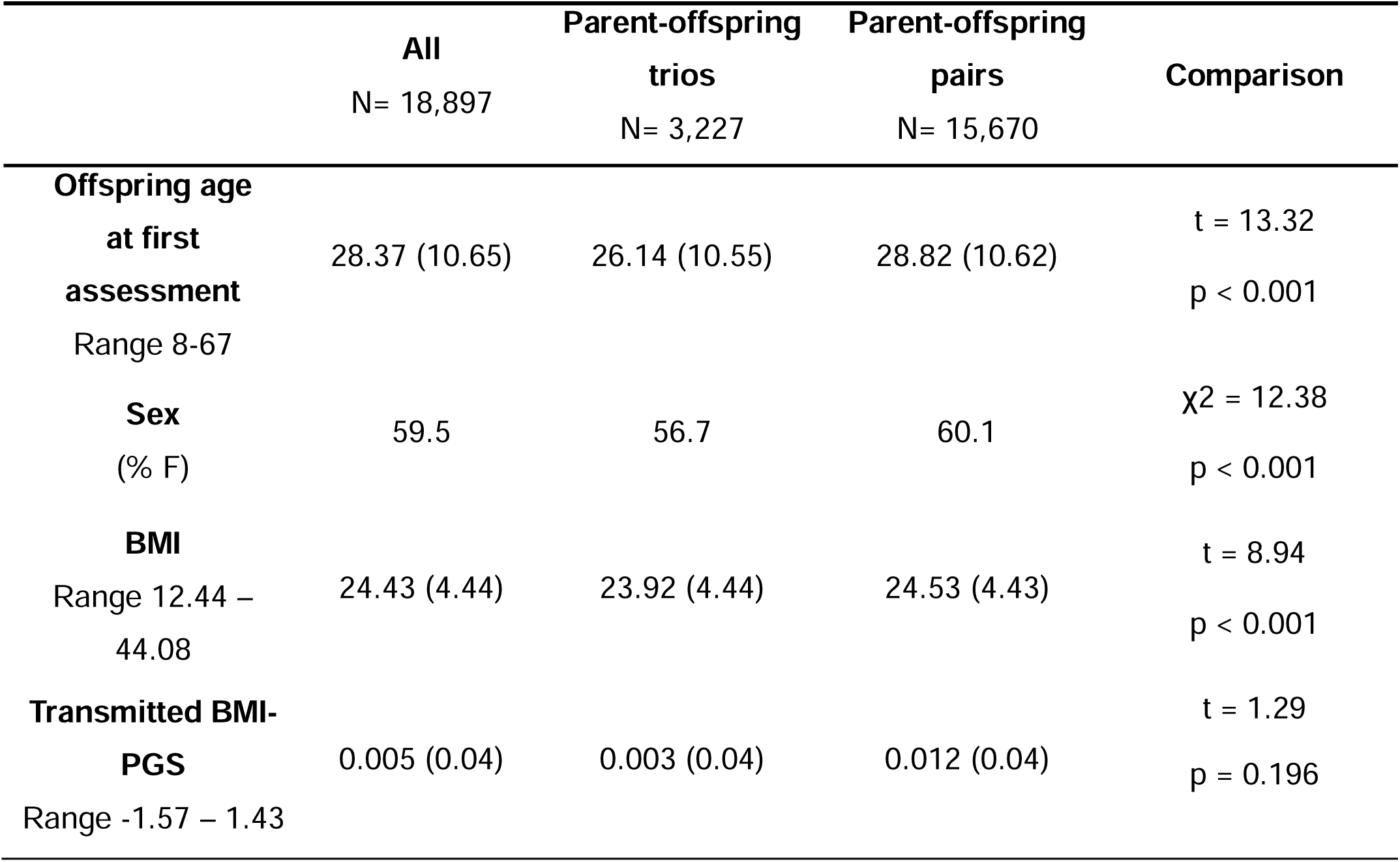
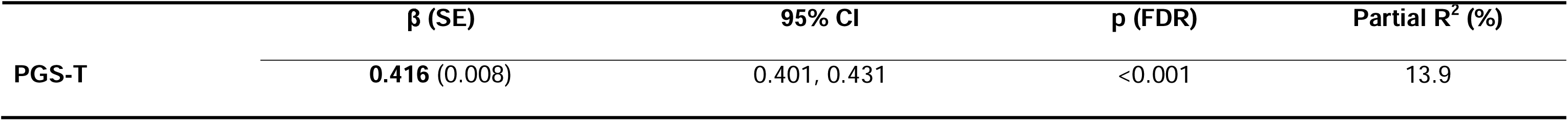
Sample characteristics of the study, presented for the total sample and stratified by parent-offspring trios vs. pairs. Comparison statistics (t-tests and chi-square (χ2) test) assess differences between parent-offspring trios and pairs. Values represent mean (standard deviation) unless otherwise specified. BMI: Body mass index; PGS: Polygenic score.

The correlation between maternal and paternal BMI-PGS was r=0.061 (p<0.001), pointing to a small degree of assortative mating in the parental generation. However, as PGSs capture only a portion of trait heritability, this correlation likely underestimates true genetic similarity between partners due to assortative mating, and some residual confounding of PGS-NT estimates cannot be ruled out.^33^

### Model comparisons

The best-fitting model included interactions (AIC= 74205.93, R²= 45.5%). In comparison, the model with main effects only had a higher AIC (74476.89) and a slightly lower R² (44.8%). Standardized effects for the model along with age and sex interactions are shown in **Table 2**.

**Table 2.**
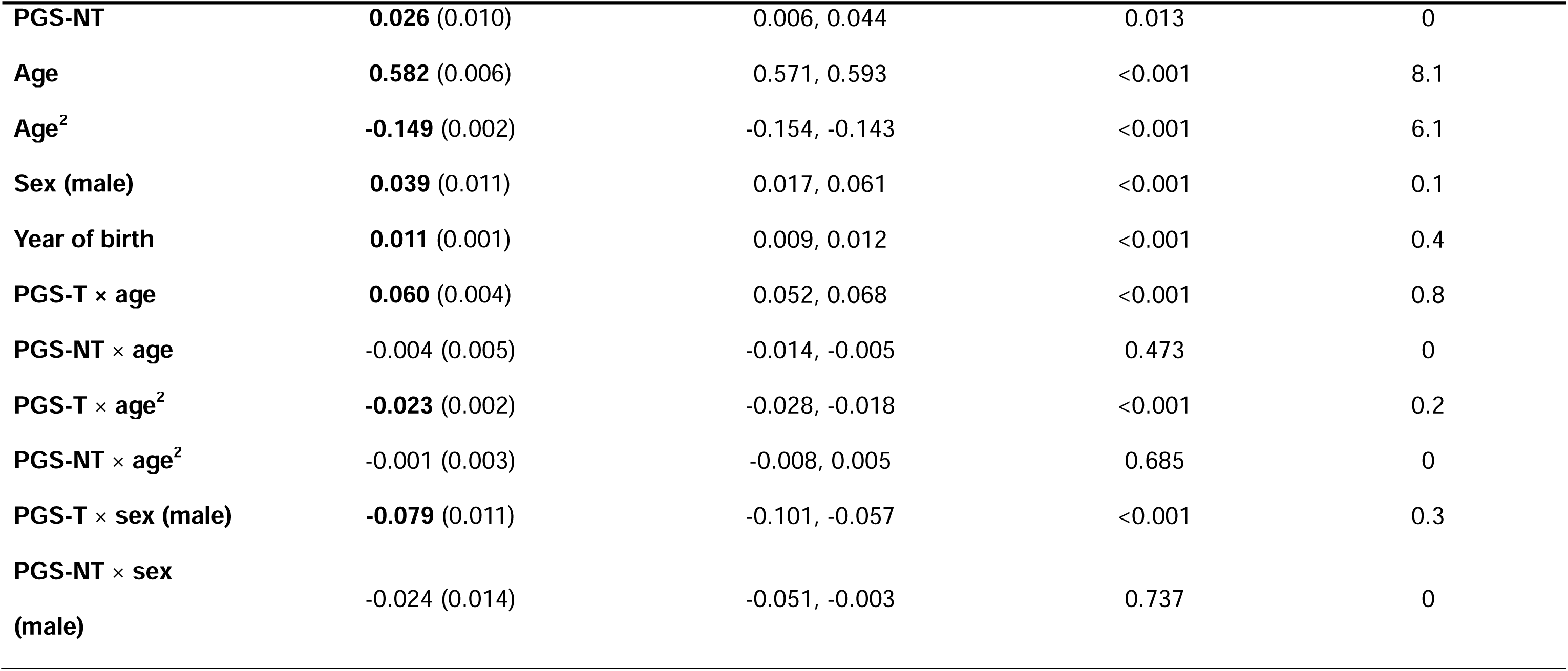
Standardized effects of model with interactions on offspring BMI. Effect sizes (β) are reported per 1 standard deviation (SD) increase in the predictor variable with standard errors (SE), 95% confidence intervals (CI), false discovery rate-adjusted p-values (p(FDR)), and percentage explained variance (Partial R^2^). Significant effects (p(FDR) < 0.05) are in bold. Non-significant terms after FDR correction: PGS-NT × age, PGS-NT × age², and PGS-NT × sex (male). Reference class for sex is female.

### Main Effects

PGS-T had a large and significant effect on offspring BMI (β=0.416, 95% CI 0.401 – 0.431, p<0.001). This corresponds to an increase of 1.85 kg/m² in BMI for each standard deviation increase in PGS-T. PGS-NT was also significantly associated with offspring BMI, though with a much smaller effect size (β=0.026, 95% CI 0.006 – 0.044, p=0.013) (**Figure 1**, **Table 2**). The ratio of PGS-NT to (PGS-T – PGS-NT) was 6.6%, providing an estimate of genetic nurture as a proportion of direct transmission; note that this ratio assumes PGS-T and PGS-NT are independent, which may not fully hold under assortative mating. Parent-of-origin analyses revealed that significant PGS-NT effects were only found for maternal PGS-NT; estimated effects for paternal PGS-NT were negligible and not significant (**Table 3**). To formally test whether these effects differed, we ran a joint model in the trio-only subset (N=3,226 offspring): maternal PGS-NT showed a significant effect (β = 0.085, SE = 0.026), paternal PGS-NT showed a null effect (β = −0.001, SE = 0.026), and the difference was statistically significant (Wald test: χ²(1) = 5.11, p = 0.024).

**Figure 1.**
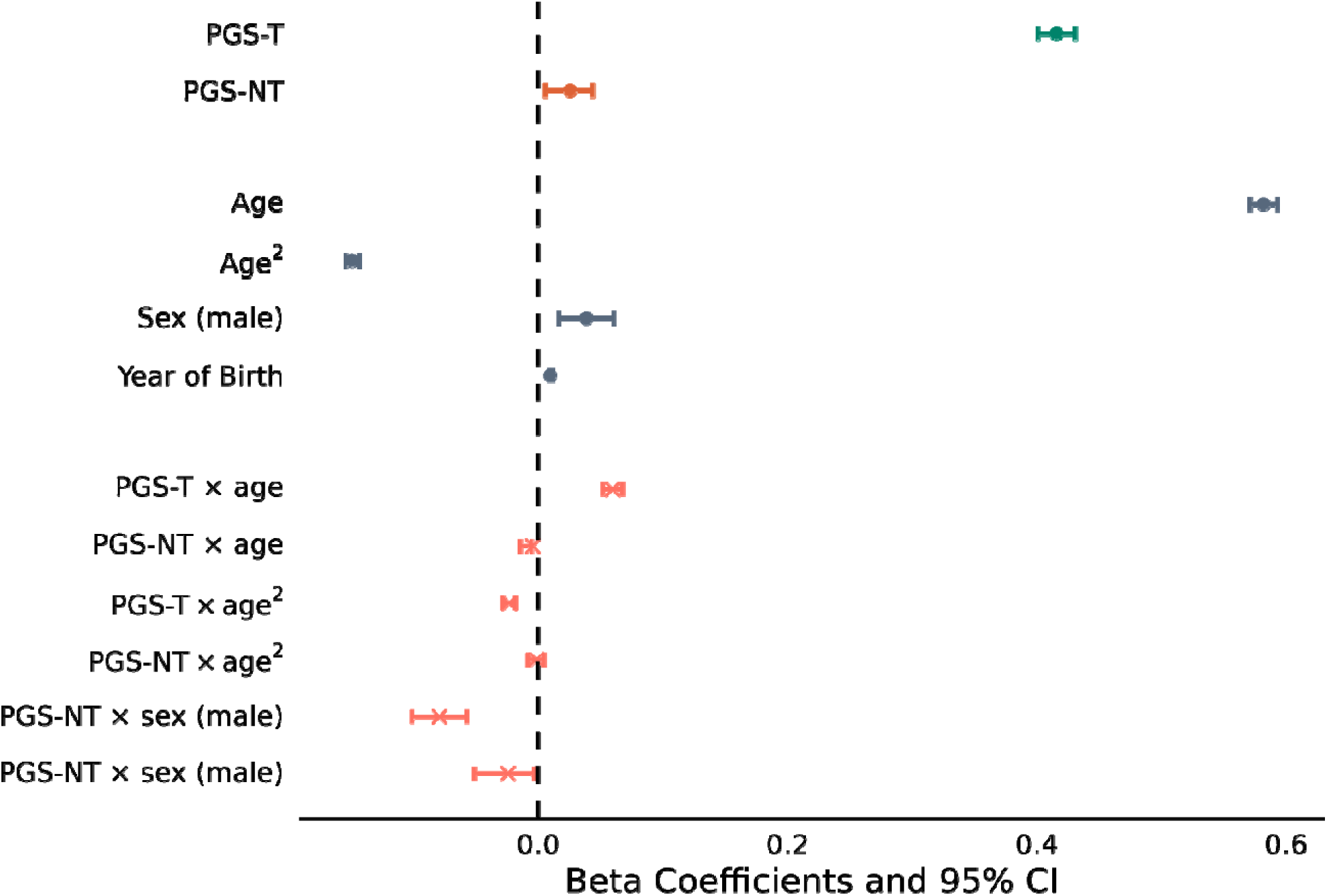
Forest plot of standardized effect sizes and 95% confidence intervals (CI) from the model predictors on offspring BMI. Blue circles represent the effects of transmitted polygenic scores (PGS-T), orange circles represent non-transmitted polygenic scores (PGS-NT), grey circles correspond to covariates, and pink crosses indicate interaction terms. Error bars represent 95% confidence intervals. Reference class for sex is female.

**Table 3.** Effects of transmitted (PGS-T) and non-transmitted (PGS-NT) polygenic scores for BMI, separated by maternal and paternal haplotypes. Effect sizes ( β) are reported per 1 standard deviation (SD) increase in PGS with standard errors (SE), 95% confidence intervals (CI), and false discovery rate-adjusted p-values (p(FDR)). Significant effects (p(FDR) < 0.05) are in bold. Nmaternal = 12,977; Npaternal = 9,141.

|  | Maternal |  |  | Paternal |  |  |
| --- | --- | --- | --- | --- | --- | --- |
| | $\beta$ (SE) | 95% CI | p (FDR) | $\beta$ (SE) | 95% CI | p (FDR) |
| **PGS-T** | **0.355** (0.010) | 0.335,0.375 | <0.001 | **0.347** (0.011) | 0.325,0.369 | <0.001 |
| **PGS-NT** | **0.030** (0.010) | 0.010, 0.050 | 0.004 | 0.003 (0.011) | -0.019,0.025 | 0.737 |

### Interaction effects

The PGS-T showed significant interactions with age, age^2^ and sex (**Table 1**). The interaction with age^2^ was negative (β=-0.023, 95% CI −0.028 – −0.018, p<0.001), indicating that the effect of PGS-T on offspring BMI leveled off across age and plateaued after approximately the age of 50 (**Figure 2**). For PGS-NT, the interactions with age and age^2^ were not statistically significant, suggesting that the genetic nurture effects on BMI remained consistent across age (**Figure 2**). Because the age range of the sample (8–67 years) far exceeds the follow-up period (∼10 years), age-related patterns in genetic effects primarily reflect cross-sectional differences between individuals rather than within-person change over time. The negative interaction with sex (β = –0.079, 95% CI –0.101 to –0.057, p<0.001) indicated a weaker direct genetic effect in males compared to females.

**Figure 2.**
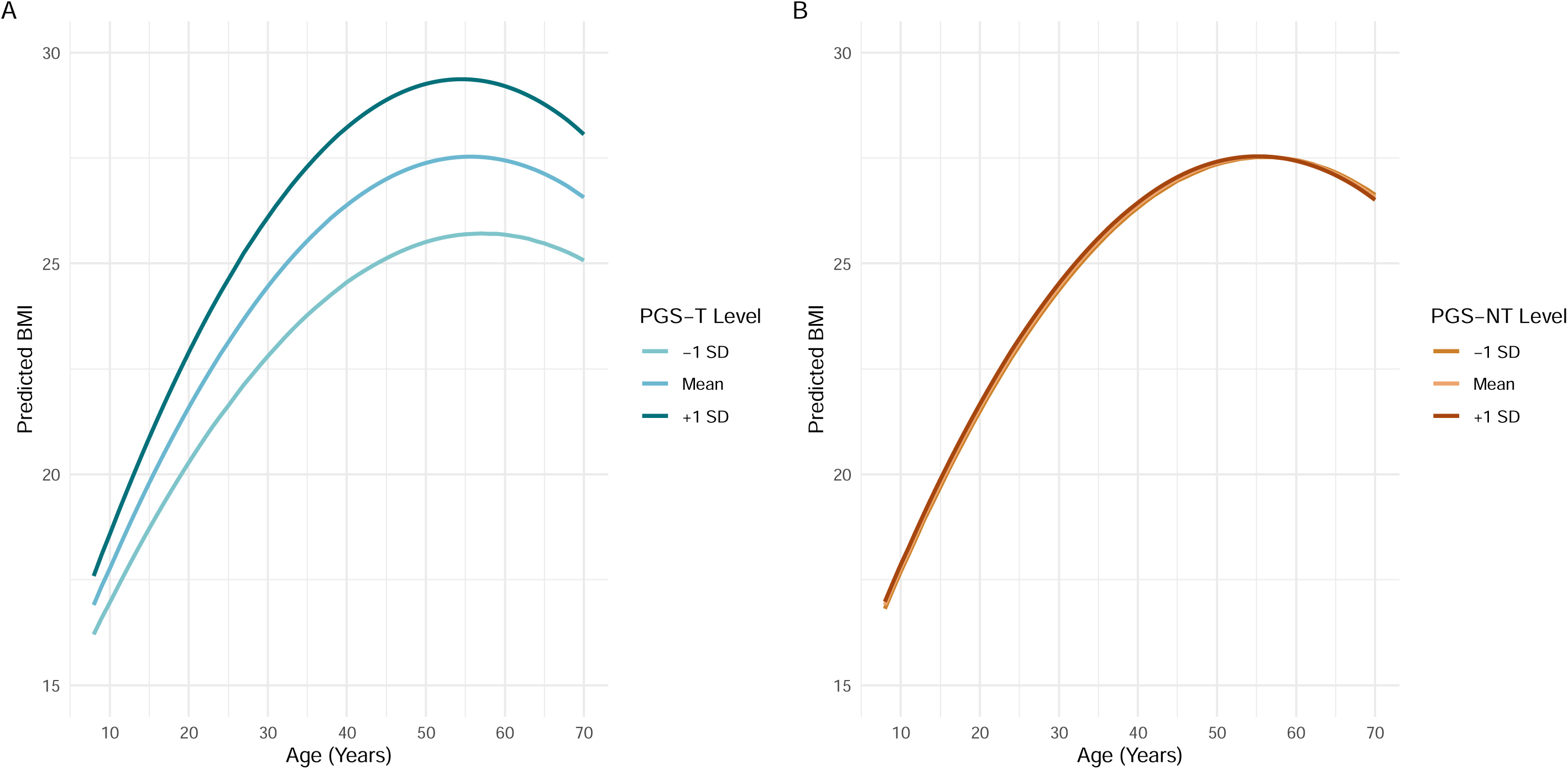
Predicted offspring BMI across age by levels of transmitted (PGS-T; panel A) and non-transmitted (PGS-NT; panel B) polygenic scores for BMI. Predictions are based on a linear mixed-effects model including age and age² interactions with both polygenic scores with all covariates (sex, year of birth, genotyping array) held constant. The shaded area represents 95% confidence intervals.

### Sensitivity analyses

Analyses based on parent-offspring trios only (N= 3,227) yielded similar results as our main model (PGS-T β=0.389, 95% CI 0.353 – 0.424, p<0.001; PGS-NT β=0.042, 95% CI 0.007 – 0.076, p=0.017) (**Table S1**), indicating that results of the full sample including parent-offspring pairs are robust. The slightly larger PGS-NT estimate in the trio-only analysis (β=0.042 vs. β=0.026 in the full sample) likely reflects attenuation bias: imputed non-transmitted alleles in parent-offspring pairs introduce measurement error that shrinks estimates toward the null. The confidence intervals of the two estimates overlap, indicating the difference is not statistically significant. Results of age-stratified analyses were concordant with our main analyses (**Table S2**). GAMM results were in line with the main model but suggested that the effect of PGS-T stabilizes instead of declining in older adulthood (**Figure 3**).

**Figure 3.**
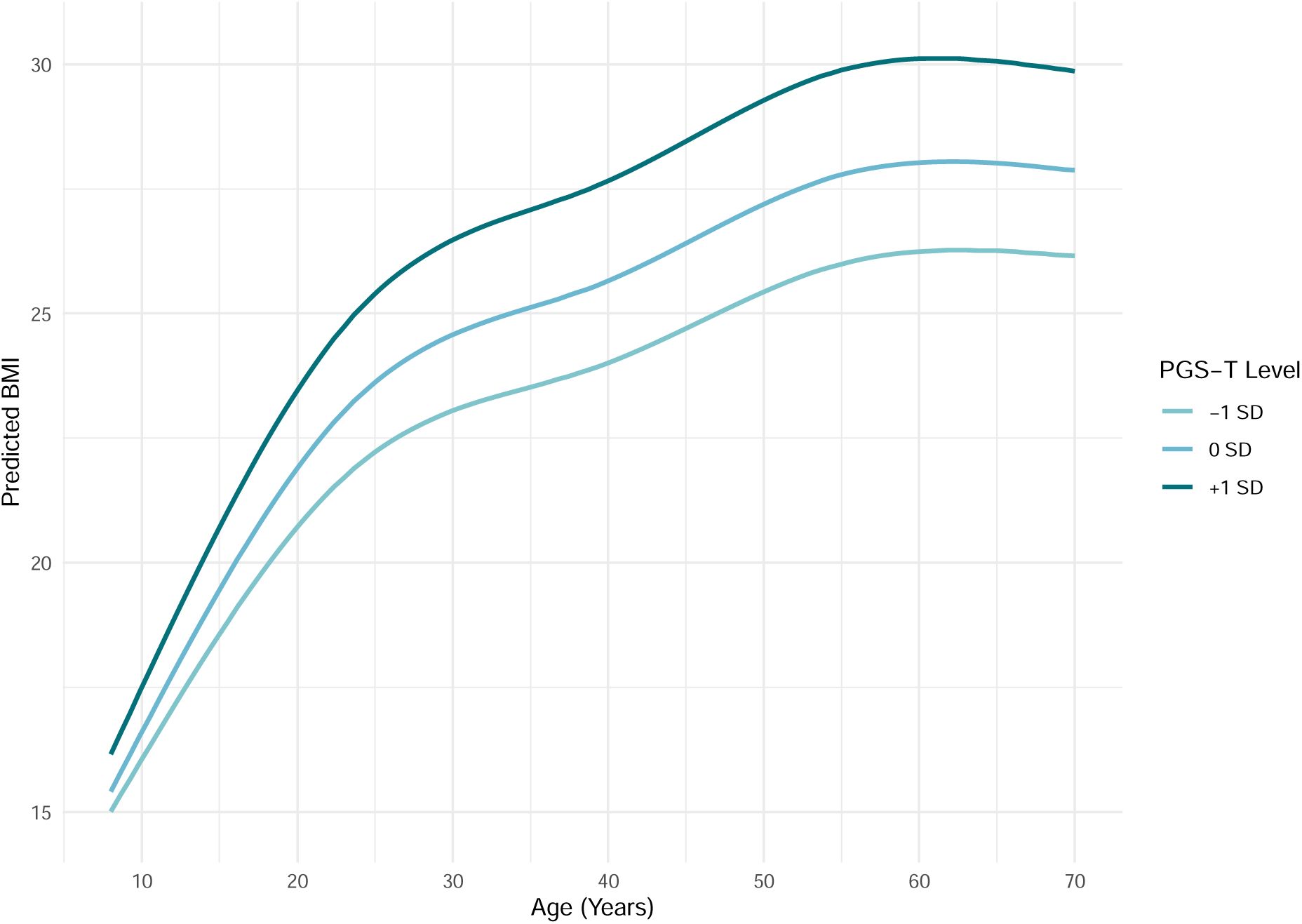
Predicted BMI across age based on a generalized additive mixed model (GAMM) including a smooth interaction between age and transmitted polygenic scores for BMI (PGS-T). Predictions are shown for the mean and ±1 SD levels of PGS-T, with covariates (sex, year of birth, genotyping array, and PGS-NT) held constant. Shaded areas represent 95% confidence intervals.

## Discussion

In this study, we used data from a large general population cohort with repeated BMI measurements across age to investigate direct transmission and genetic nurture effects on BMI. We leveraged a powerful multi-ancestry BMI-PGS and a novel haplotyped based method to generate PGS-T and PGS-NT. Our findings showed a large effect of direct genetic transmission and a small genetic nurture effect on offspring BMI, with the latter suggesting limited environmental influences of parental genotypes associated with BMI on offspring’s BMI. The PGS-NT effect was found only for the maternal haplotype, pointing to a differential contribution of maternal and paternal genes on offspring BMI. Interactions of PGS-T with age indicated a non-linear relationship, in which the direct effects initially increased from childhood to adolescence and tended to stabilize after approximately age 55. In contrast, the impact of PGS-NT was not moderated by age in our main analyses. Below, we discuss these findings in more detail.

### Direct transmission and genetic nurture effects

We found a large effect of PGS-T, which explained the greatest proportion of variance in our main model (*R*^2^=13.9%), consistent with expectations for European ancestry populations based on the BMI-PGS development paper (*R*^2^ between 12.9% and 17.6%)^14^. This strong direct genetic effect reflects the use of the most predictive BMI-PGS to date, derived from a GWAS meta-analysis that included over 5.1 million individuals from diverse populations. The methodological strengths of using a powerful PRS in combination with a large parent-offspring sample positioned our study better to detect genetic nurture effects than previous studies. Although we observed a significant PGS-NT effect, it was small, with the effect 6.6% of the direct transmission estimate. Our results are consistent with parental BMI-associated alleles having limited influence on offspring BMI through non-transmitted pathways, though we cannot exclude contributions from assortative mating, dynastic effects, or prenatal mechanisms.

Our finding of small genetic nurture effects on BMI clarifies the mixed results in prior literature. When effects are subtle, weak polygenic scores and small sample sizes will consistently fail to detect them^9,10,23,24^. More recently, two studies showed results that are broadly consistent with the current study. Ghatan et al. (2025), using a joint regression of child’s, mother’s, and father’s PGS in Generation R (N=4,488, ages 6–13), found significant genetic nurture effects at ages 9 and 13 (β=0.05, 95% CI: 0.01–0.09), with no significant maternal versus paternal difference. Wright et al. (2025), using an instrumental variable approach in the UK Millennium Cohort Study (N=2,630, ages 3–17), found significant maternal but not paternal indirect effects from age 11 onwards. With the largest sample to date, repeated measures spanning childhood through late adulthood, a PGS derived from the largest BMI GWAS available, and a design that directly compares maternal versus paternal genetic nurture, our study provides the strongest test of this question to date, showing that genetic nurture effects on BMI are small and driven by the maternal side.

### Maternal versus paternal effects

Our results showed a significant PGS-NT effect in the maternal model but not in the paternal model. This finding aligns with previous studies which also suggest that maternal genetic nurture effects may play a more prominent role in BMI than paternal ones ^13^. Several explanations could underlie this maternal dominance, including a greater maternal role in shaping offspring dietary habits and overall rearing environment^25^. Another possible explanation is the fetal overnutrition hypothesis, which proposes that maternal adiposity during pregnancy contributes to higher offspring obesity risk^26,27^. While the mechanisms remain unclear and require further investigation, these findings reinforce the conclusion that parent-of-origin analyses are an important first step in unraveling these mechanisms, although power challenges related to the smaller availability of genotype data for fathers persist.

### Influence of direct transmission and genetic nurture across age

Our results show that the effect of PGS-T on BMI increases between childhood and early adulthood, but levels off after middle adulthood. Our main model captured this trend with the quadratic term for age, and the GAMM analyses further refined it, showing that rather than declining, the effect stabilizes over time. The relationship we observed is consistent with a prior study by Song and colleagues, which found that PGS-T effects on BMI increased from early to middle adulthood and peaked there, before declining in later life^21^. Pooled twin analyses also support this pattern by showing rising heritability of BMI from childhood to adolescence, followed by a decline in older adulthood due to increasing unique environmental influences^8,28^. Despite differences in sample age and methods, the general age-dependent genetic influence on BMI observed in our results is hence concordant with previous studies. However, the interaction effect is relatively small, suggesting that that while the influence of PGS-T on BMI varies across age, this age-dependent variation itself contributes only modestly to overall BMI differences in our sample.

Our findings of stabilizing genetic transmission effects in middle and older adulthood also align with previous work demonstrating high genetic correlations (0.89-1.00) for BMI between ages 40-73 years^29^. Their genomic structural equation modeling showed that BMI variation across this age range is underpinned by a stable set of genetic influences rather than age-specific genetic factors. Our study extends these insights by capturing the full trajectory from childhood through older adulthood, revealing the strengthening of genetic effects before the period of stability observed in Gillespie’s study.

Theoretical models suggest that genetic nurture should be stronger during childhood and adolescence, when parents and offspring share a household, than in adulthood^30^. Our results do not support this expectation: we found no statistically significant interaction between PGS-NT and age, suggesting that genetic nurture effects on BMI may remain stable across the lifespan. However, given the small size of the PGS-NT effect, our power to detect subtle interactions remains limited. Previous studies on two separate cohorts with longitudinal data from the UK^12,13^ point to an increasing impact of genetic nurture effects on BMI over time; however, these studies only included data up to age 17. The increase observed before age 17 may reflect the growing influence of the rearing environment from childhood to adolescence, a period when environmental exposures accumulate. Overall, our findings suggest a persistent, albeit small, impact of genetic nurture on BMI across age. Using repeated measures data across a large age range, as in our study, can be informative about the long-term impact of genetic nurture effects on health outcomes, which can be overlooked in studies that include a more limited age range or a single assessment.

### Moderation by sex

Our results indicate a significant negative interaction between PGS-T and male sex, suggesting a weaker effect of offspring’s own PGS on BMI in males compared to females. While evidence exists for sex-specific genetic factors underlying BMI^8^, which could reasonably lead to variation in PGS performance across sexes, our findings contrast with those of the BMI-PGS development study, which reported slightly higher variance explained by the PGS in males (17.9%) than in females (17.3%) in the UK Biobank^14^. The minimal sex differences observed in both studies, alongside the small interaction effect in our study, suggest that these differences may be more reflective of cohort-specific characteristics than of any inherent difference in the PGS’s ability to capture genetic influences across sexes. Sex-stratified analyses confirmed that PGS-T explained slightly more variance in females (partial R²=16.9%) than in males (partial R²=15.9%), consistent with the interaction term (see Supplementary Table S3). This contrasts with the pattern reported in the PGS development sample, where variance explained was marginally higher in males (17.9% vs. 17.3%; Smit et al., 2025), though the direction in our data is directly consistent with the significant PGS-T × sex interaction observed in our model.

### Implications and future research directions

Ultimately, our findings indicate a persistent yet small impact of genetic nurture on BMI across age. While the rearing environment remains important for offspring BMI and could be targeted by interventions such as school-based nutrition programs^31^, parental BMI-associated alleles seem to have only limited influence on this environment. Other parental genetic environmental factors may play more substantial roles. Future studies could extend our findings by examining genetic nurture through alternative genetic pathways. A promising example is educational attainment, which was shown to have a genetic nurture effect on BMI in that parents with a higher PGS-NT for educational attainment had offspring with lower BMI^7^. Furthermore, future research could extend and deepen the evidence for a maternally driven genetic nurture effect by genetically informed studies incorporating prenatal factors, birth weight, parenting, and home environment to elucidate maternal-specific pathways.

This study is the first to examine genetic nurture effects across such a wide age range, spanning childhood through late adulthood. Our findings suggest that these effects remain stable rather than diminish over time, challenging the expectation that genetic nurture wanes after adolescence. However, it remains unclear whether this pattern extends to other traits, and most studies conducted so far have focused on children and adolescents. Applying similar methods to other traits and including older cohorts could provide further insight into the long-term role of the rearing environment through genetic nurture.

### Limitations

Despite the several strengths of this study, there are limitations that should be considered when interpreting the findings. First, although broad population stratification was corrected for, and the Lifelines subsample consists of a relatively homogeneous population, a small correlation between parental BMI-PGS was observed. Given that PGSs capture only a portion of trait heritability, this correlation likely underestimates true genetic similarity between partners,^33^ and residual confounding from assortative mating and subtle population structure may remain and could influence the PGS-NT estimates^32^ However, the maternal-specific pattern observed in our parent-of-origin analyses argues against AM as the primary source of this association, as AM would be expected to inflate both maternal and paternal estimates equally. Furthermore, the generalizability of our findings is limited, as both cohorts used in our analyses were of European ancestry and do not represent diverse populations. Finally, although our dataset includes repeated BMI measurements across a wide age range, the design is not purely longitudinal. Because the age range of the sample (8–67 years) far exceeds the follow-up period (∼10 years), age-related patterns in genetic effects primarily reflect cross-sectional differences between individuals rather than within-person change over time.

### Conclusions

Our findings reveal distinct roles of direct genetic transmission and genetic nurture in shaping BMI across age. Direct genetic effects were strong, increasing from childhood to adulthood before stabilizing around age 50, and explained 13.9% of BMI variance. In contrast, genetic nurture effects were small (only 6.6% of the direct transmission effect), were primarily driven by maternal PGS-NT, and showed no clear age-dependent pattern, suggesting they may operate consistently across the lifespan. By leveraging repeated measures across a broad age range, this study improves our understanding of how parental genetic influence unfolds over time and reinforces the value of incorporating parent-of-origin models in intergenerational research.

## Methods

### Sample

Lifelines is a multi-disciplinary prospective population-based cohort study examining, in a multi-generational design, the health and health-related behaviors of 167,729 persons living in the North of the Netherlands. It employs a broad range of investigative procedures in assessing the biomedical, socio-demographic, behavioral, physical and psychological factors that contribute to the health and disease of the general population, with a special focus on multi-morbidity and complex genetics. All participants provided written informed consent. The Lifelines Cohort Study was approved by the Medical Ethics Committee of the University Medical Center Groningen in The Netherlands^16,17^.

Participants were recruited in different ways: initially via general practitioner invitations, followed by family member recruiting, and independent registrations through the Lifelines website. Data were collected across three main assessment waves: the baseline (2007–2013), the second wave (2014–2017), and the third wave (2019–2023). BMI was calculated using height and weight measurements obtained on-site during each wave, following standardized anthropometry protocols.

The study sample consisted of 18,897 genotyped offspring with at least one genotyped parent in Lifelines. To minimize the impact of measurement errors or outliers that could bias the analyses, BMI measurements were excluded if they met any of the following criteria: self-reported diagnosis of an eating disorder, pregnancy at the time of BMI measurement, or extreme BMI values (>±4 standard deviations from the age-adjusted mean).

### Genotype data

Imputed genetic data was available for 79,988 Lifelines participants genotyped across three batches. Quality control (QC) of markers and samples was performed separately per batch.

The pre-imputation QC criteria are described in detail in Supplementary Information. In brief, markers with a high missing rate or low minor allele frequency, markers that were duplicated or monomorphic, and markers that deviated significantly from Hardy-Weinberg equilibrium were removed. Post QC data from each array was imputed using the Sanger Imputation Service with the Haplotype Reference Consortium v1 as reference panel. To get a common set of markers for parent and offspring genotyped in any array, we selected an overlap of imputed markers with imputation quality scores equal to or above 0.8 across arrays. Samples that had a high missing rate, heterozygosity outliers, or samples that were identified as mix-ups were filtered out. Samples were restricted to European ancestry based on a principal component analysis using the 1000 Genomes reference to avoid confounding due to broad population stratification. In cases of individuals genotyped in more than one array, data from the most recent release was used.

### Non-transmitted alleles inference

To partition direct genetic effects from genetic nurture effects, we implemented a computational phasing approach to identify accurately which parental alleles were transmitted to offspring and which remained non-transmitted, using our validated haplotype-based method from our previous study of genetic nurture effects on educational attainment^15^. In short, we used the method SHAPEIT5^18^ to estimate haplotypes including pedigree information. Offspring haplotypes were then compared to parental haplotypes using tiles of 150 adjacent markers on each chromosome. The best match between the parent and offspring tiles, taking recombination spots into account, was used to determine which parental tiles were transmitted to the offspring. The remaining non-transmitted alleles were recorded in a separate dataset, and for parent-offspring pairs, the non-transmitted alleles of the unobserved parent were set as missing. We validated our method by comparison with standard software^19^ in parent-offspring trios and found a concordance rate for the non-transmitted alleles of 99.8%. Furthermore, the identification of non-transmitted alleles was confirmed to be unaffected by missing parental data through simulations of pairs from trios.

### Polygenic scores

To compute PGSs for BMI, we utilized weights from the latest polygenic score for BMI, derived from GIANT+23andMe GWAS summary statistics based on more than 5 million individuals^14^. In brief, the GWAS meta-analysis included 23andMe and excluded UK Biobank, ALSPAC, and BioMe to allow for independent validation. PRS-CSx^20^ was used to generate ancestry-specific effect estimates, which were meta-analyzed to produce a single set of posterior weights. These weights are designed to be applicable across diverse target populations, reducing the need for population-specific linear combinations of ancestry-specific subscores. We used the ‘meta’ PGS, which combines effect estimates from five ancestries into a single PGS. In our study, the PGS included 964,006 markers that overlapped with the Lifelines genotype data.

### Statistical models

We used linear mixed-effects models to assess the relationships between PGS for BMI based on transmitted (PGS-T) and non-transmitted (PGS-NT) parental haplotypes on offspring BMI. Random intercepts were included to account for the hierarchical structure of the data, specifically the repeated BMI measurements within individuals (offspring ID) and the correlation among sibling relationships in the data (family ID). We tested the interactions between the PGSs and age (both linear and quadratic) to investigate if the influence of the PGS-T and PGS-NT changes with age. Given previous evidence^21^, we did not expect the relationships between PGS-T, PGS-NT and BMI to exhibit complex nonlinear patterns with multiple peaks or troughs, and therefore deemed this relatively simple, straightforward approach appropriate to capturing general nonlinear age-related trends.

To investigate parent-of-origin effects, we partitioned the PGS-T and PGS-NT into maternal and paternal haplotypes for separate analyses including only maternal or paternal PGSs. Genotyping array and year of birth were included as covariates in all models. To control for residual population stratification, the first 10 genetic principal components were regressed out of all PGS prior to analysis. We compared models with and without interaction terms to assess whether including these interactions significantly improved model fit with likelihood ratio tests and AIC (Akaike Information Criterion).

We applied false-discovery rate (FDR) correction across the tests: 10 tests for the main effects (PGS-T and PGS-NT on BMI), interactions with age and age^2^ (PGS-T × age, PGS-T × age², PGS-NT × age, PGS-NT × age² on BMI), and analyses split by maternal/paternal PGS (maternal PGS-T, maternal PGS-NT, paternal PGS-T, paternal PGS-NT on BMI).

To assess assortative mating in the parental generation of our sample, we computed the Pearson correlation between maternal and paternal PGSs for BMI in the parent-offspring trios. High assortative mating for a trait can confound genetic nurture effects detected with a PGS-NT.

We preregistered this study on OSF (number 9YFCM), with the main deviations concerning the BMI outlier threshold (changed from ≥3 SD to ≥4 SD from the age-adjusted mean), the addition of sex interactions and year of birth as covariates, and the postponement of the pre-registered meta-analysis; full details are provided in the Supplementary Information.

### Sensitivity analyses

To ensure the robustness of our findings, we conducted various sensitivity analyses. First, we restricted the sample to parent-offspring trios only to minimize estimation errors that can arise when imputing PGS-NT in parent-offspring pairs where part of the genetic data is missing for one parent. Second, we stratified the data by age groups to examine the age-related effects that might be obscured in our main model, given the relatively low number of BMI measurements in the childhood and late adulthood groups compared to young and middle adulthood (19-50 years old). This imbalance could potentially skew the overall estimates toward associations reflecting adulthood measurements, making it harder to detect variation in earlier or later life stages. Lastly, we fitted generalized additive mixed models (GAMM) to explore potential more complex, non-linear associations between PGS-T and age without constraining the analysis to a predetermined quadratic pattern. Full details of these analyses are provided in the Supplementary Information.

## Data availability

Data may be obtained from a third party and are not publicly available. Researchers can apply to use the Lifelines data used in this study. More information about how to request Lifelines data and the conditions of use can be found on their website (https://www.lifelines.nl/researcher/how-to-apply).

## Code availability

The R analysis scripts supporting this study are publicly available as a GitHub Gist: https://gist.github.com/vtpons/470ee86bee041237410b48d2f6466c20 (BMI_PGS_models.R).

## Ethics statement

The Lifelines protocol has been approved by the UMCG Medical ethical committee under number 2007/152.

## Acknowledgements

This work was supported by grants from the National Institute of Mental Health (NIMH grant R01MH125902 and R01DA052453). HvL was supported by a VENI grant from the Talent Program of the Netherlands Organization of Scientific Research (NWO-ZonMW 09150161810021). The Lifelines initiative has been made possible by subsidy from the Dutch Ministry of Health, Welfare and Sport, the Dutch Ministry of Economic Affairs, the University Medical Center Groningen (UMCG), Groningen University and the Provinces in the North of the Netherlands (Drenthe, Friesland, Groningen). We acknowledge the services of the Lifelines Cohort Study, the contributing research centers delivering data to Lifelines, and all the study participants.

