## Supplementary Information for "Genetic nurture and direct genetic transmission effects on body mass index across age"

### 1. Exclusion criteria

To minimize the impact of measurement errors or outliers, BMI measurements were excluded based on specific criteria. A total of 299 individuals were excluded because they self-reported having an eating disorder at wave 1. Then, 45 BMI measurements were excluded as they occurred within 9 months of the individual reporting being pregnant. Finally, 173 BMI measurements were excluded for being more than 4 standard deviations away from the age-adjusted mean.

### 2. Deviations from pre-registration

Deviations from the pre-registered analysis plan (<https://osf.io/9yfcm>) were made to improve the validity of the analyses and to align the approach with the data and previous evidence.

1. The BMI outlier threshold was changed from ≥3 SD to ≥4 SD from the age-specific mean to avoid removing valid observations; the ≥3 SD threshold excluded 420 plausible BMI measurements.
2. Interactions between PGS-T and PGS-NT with sex were included in the main model, although not specified in the preregistration, because previous work showed that the predictive performance of BMI-PGS differs by sex^14^.
3. Year of birth was added as a covariate to account for cohort effects influencing genetic effects on BMI^22^.
4. The preregistered meta-anlysis was postponed because leave-one-out summary statistics were not available for the QIMR cohort at the time of analysis. We also consider including a larger set of cohorts for a meta-analysis in the future to increase the generalizability of the results.

### 3. BMI distribution

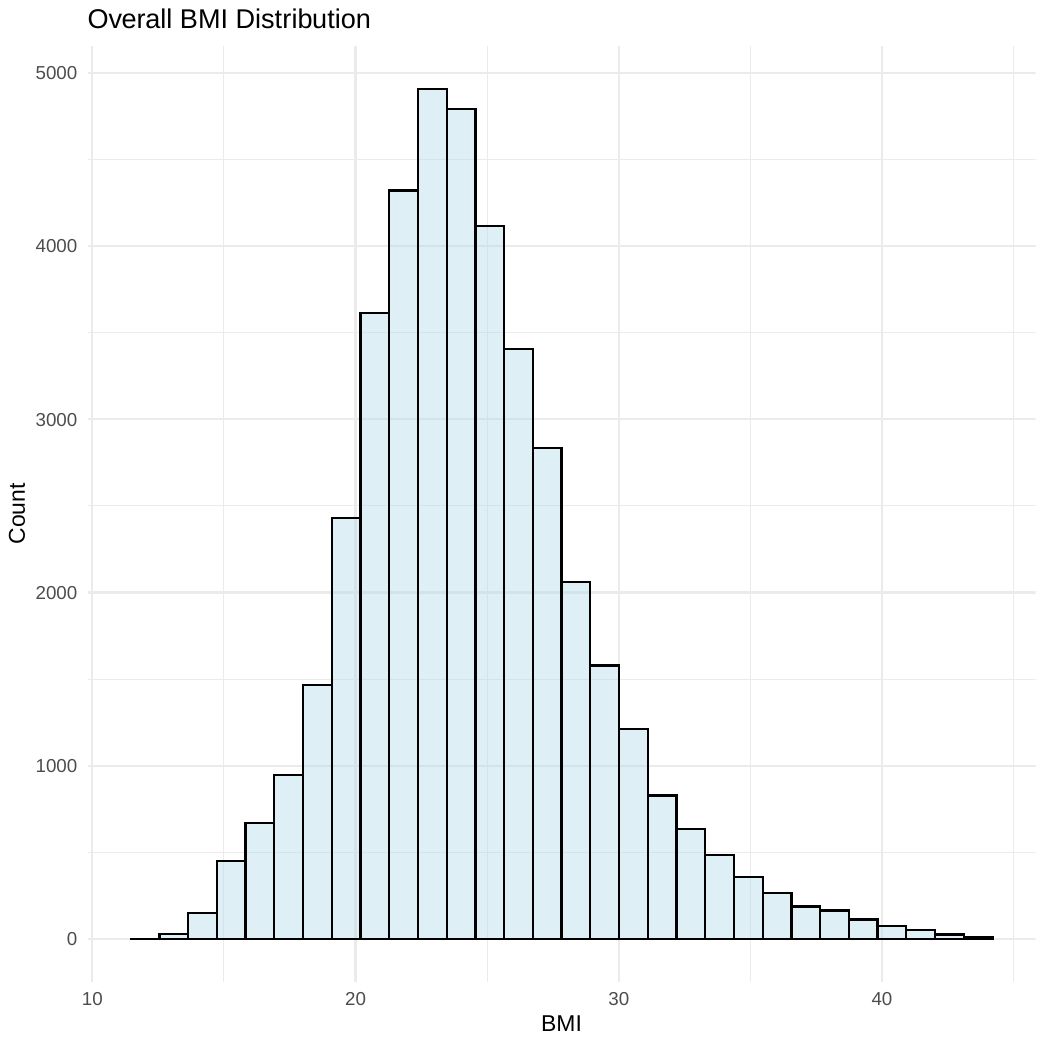
**Figure 1.** Histogram of BMI distribution in Lifelines sample (N= 18,897) after applying excluding criteria.

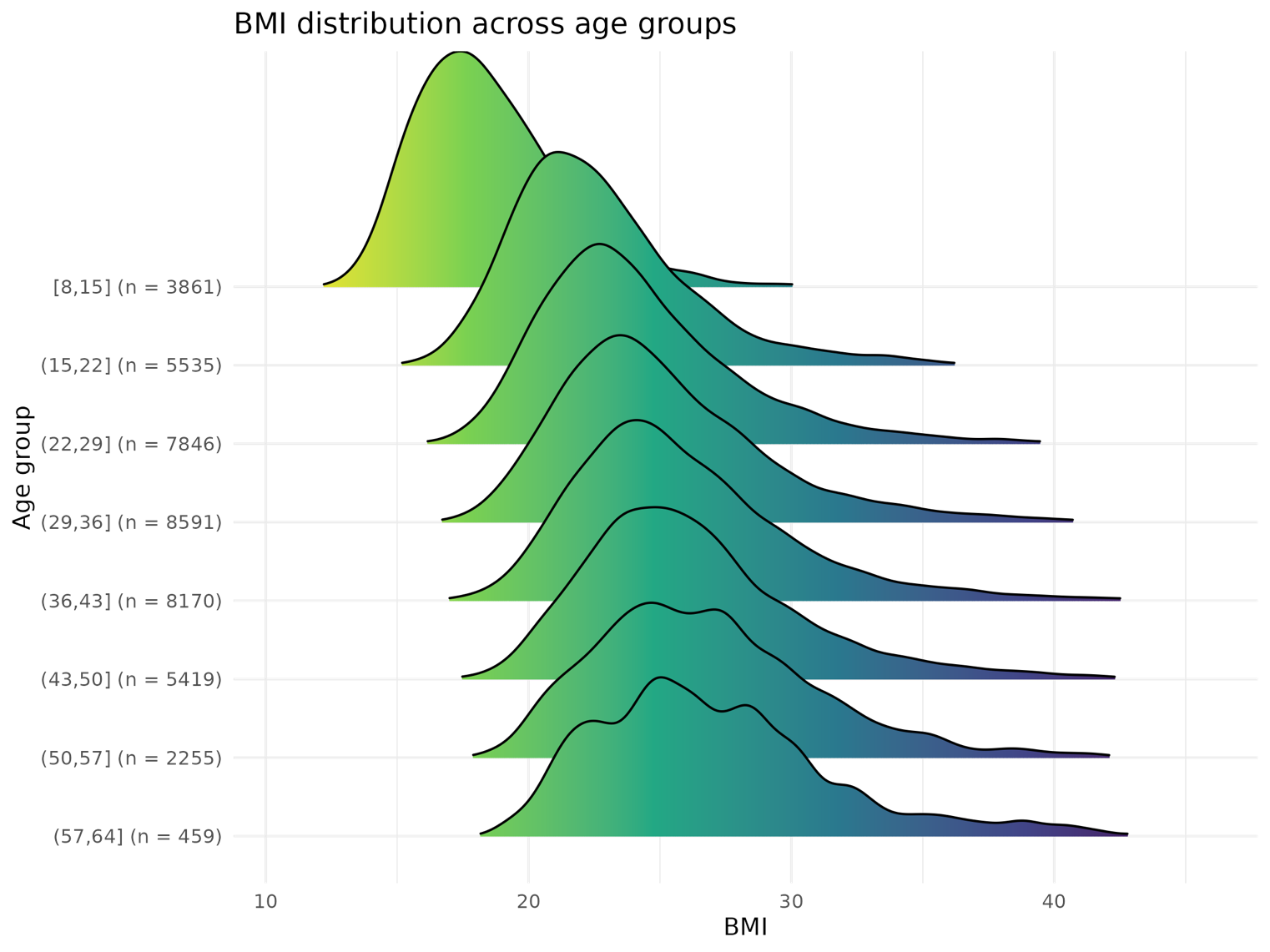
**Figure 2.** BMI distribution across age groups in the Lifelines sample (N= 18,897). Each curve represents the smoothed density of BMI values within a 7-year age group, with color indicating BMI values. Curves for the oldest age groups are more irregular due to the smaller number of available BMI measurements (n).

### 4. Analyses restricted to parent-offspring trios

|  | **β (SE)** | **95% CI** | **p** |
| --- | --- | --- | --- |
| **PGS-T** | **0.389** (0.018) | 0.352, 0.424 | <0.001 |
| **PGS-NT** | **0.042** (0.017) | 0.007, 0.076 | 0.017 |
| **Age** | **0.580** (0.012) | 0.555, 0.604 | <0.001 |
| **Age^2^** | **-0.176** (0.006) | -0.189, -0.164 | <0.001 |
| **Sex (male)** | **0.020** (0.025) | 0.028, 0.070 | 0.415 |
| **Year of birth** | **0.010** (0.001) | 0.007, 0.014 | <0.001 |
| **PGS-T × age** | **0.052** (0.009) | 0.033, 0.070 | <0.001 |
| **PGS-NT × age** | -0.014 (0.009) | -0.032, 0.004 | 0.135 |
| **PGS-T × age^2^** | **-0.025** (0.006) | -0.037, -0.012 | <0.001 |
| **PGS-NT × age^2^** | -0.008 (0.006) | -0.020, 0.004 | 0.218 |
| **PGS-T × sex (male)** | -0.048 (0.025) | -0.098, 0.006 | 0.053 |
| **PGS-NT × sex (male)** | -0.038 (0.024) | -0.086, 0.010 | 0.122 |

**Table S1.** Standardized effects of predictors on offspring BMI restricted to a subsample of parent-offspring trios only (N=3,227) from Lifelines. These analyses were conducted as a sensitivity check to ensure that results remain robust when no genetic data imputation is required (parent-offspring pairs). Effect sizes (β) are reported per standard deviation increase in PGS-T, with standard errors (SE), 95% confidence intervals (CI). Significant effects (p < 0.05) are in bold. Reference class for sex is female.

### 5. Analyses stratified by age groups

|  | **PGS-T** | | **PGS-NT** | |
| --- | --- | --- | --- | --- |
|  | **β (SE)** | **p** | **β (SE)** | **p** |
| **Childhood** 8-12 years old  n measurements = 1,583 | 0.652 (0.146) | <0.001 | -0.148 (0.180) | 0.410 |
| **Adolescence** 13-18 years old  n measurements= 3,813 | 0.342 (0.068) | <0.001 | -0.006 (0.083) | 0.944 |
| **Young adulthood** 19-30 years old  n measurements= 12,509 | 0.427 (0.012) | <0.001 | 0.025 (0.016) | 0.011 |
| **Middle adulthood**  31-50 years old  n measurements= 20,939 | 0.356 (0.009) | <0.001 | 0.019 (0.011) | 0.010 |
| **Late adulthood** 51-70 years old  n measurements= 2,762 | 0.266 (0.057) | <0.001 | 0.048 (0.080) | 0.547 |

**Table S2.** Age-stratified analyses of transmitted (PGS-T) and non-transmitted (PGS-NT) polygenic scores for BMI associations with offspring BMI in the Lifelines sample. Standardized effect sizes (β) for PGS-T and PGS-NT (with standard errors and p-values) are shown for each subsample restricted to the specified age ranges. For the genetic nurture effect (PGS-NT), the stratified analyses yielded estimates near zero and negative in the childhood and young adulthood age groups, most likely due to limited power in these smaller subgroups. The absence of larger estimates in the childhood and adolescence age groups further supports our conclusion that genetic nurture effects remain consistently small and stable across age.

### 6. Generalized additive mixed models

A generalized additive mixed model (GAMM) was fitted to further explore the nonlinear association between PGS-T and age in the Lifelines sample, without imposing a predetermined shape for the association. Due to the lack of evidence for age-dependent effects of PGS-NT in our linear models, we did not further model this relationship using GAMMs.

The GAMM was implemented in R using the gamm4 package (ref) specifying a tensor product smooth of age and PGS-T and accounting for gender, year of birth, array and PGS-NT. Random intercepts were included for ID and nuclear family ID:

gamm4(bmi ~ t2(age, BMI_PRS_T_FULL_Z) + gender + yob + array + BMI_PRS_NT_FULL_Z,
random = ~(1 | nuclear_fam_id) + (1 | project_id),
data = df, method = "REML")

*Results*

The smooth interaction term between age and PGS-T was significant (edf = 14.04, F = 259.6, p < 0.001) consistent with a non-linear pattern of direct transmission effects across the age range. The model explained (R^2^) 37.7% of the variance in offspring BMI.

### 7. Sex-stratified analyses

To examine whether the variance explained by the polygenic score differs by sex, we ran the main effects model separately for female and male offspring, extracting standardized effect sizes and partial R² for PGS-T and PGS-NT in each subsample.

|  | **Predictor** | **β**  **(SE)** | **95% CI** | **p** | **Partial R² (%)** |
| --- | --- | --- | --- | --- | --- |
| **Female offspring**  (n = 25,246 obs.) | PGS-T | 0.389 (0.008) | 0.374, 0.405 | <0.001 | 16.9 |
|  | PGS-NT | 0.023 (0.010) | 0.003, 0.043 | 0.022 | 0.04 |
| **Male offspring** (n = 16,985 obs.) | PGS-T | 0.303 (0.008) | 0.288, 0.318 | <0.001 | 15.9 |
|  | PGS-NT | −0.004 (0.010) | −0.024, 0.015 | 0.667 | 0.002 |

**Table S3.** Sex-stratified effects of transmitted (PGS-T) and non-transmitted (PGS-NT) polygenic scores for BMI on offspring BMI.

### 8. Assortative mating sensitivity analysis

To assess whether the maternal PGS-NT effect could be explained by inflation due to assortative mating (AM), we ran a sensitivity analysis in the complete trio subsample (N = 7,302 observations from N = 2,289 nuclear families) including both maternal and paternal PGS-NT simultaneously in a single mixed-effects model, with age and sex interaction terms consistent with the main model specification. Under AM, maternal PGS-NT is correlated with paternal PGS-NT via the shared parental genetic correlation; including both absorbs this shared variance. If the maternal effect were AM-driven, it should attenuate substantially when conditioned on paternal PGS-NT.

| **Predictor** | **β** | | **SE** | **95% CI** | **p** |
| --- | --- | --- | --- | --- | --- |
| **PGS-T** | | 0.391 | 0.018 | 0.355, 0.426 | <0.001 |
| **Maternal PGS-NT** | | 0.085 | 0.026 | 0.034, 0.136 | 0.001 |
| **Paternal PGS-NT** | | -0.001 | 0.026 | −0.050, 0.050 | 0.988 |
| **PGS-T × age** | | 0.053 | 0.009 | 0.034, 0.072 | <0.001 |
| **PGS-T × age²** | | −0.025 | 0.006 | −0.037, −0.012 | <0.001 |
| **Maternal PGS-NT × age** | | −0.004 | 0.014 | −0.031, 0.023 | 0.765 |
| **Paternal PGS-NT × age** | | −0.025 | 0.013 | −0.051, 0.001 | 0.065 |
| **PGS-T × sex (male)** | | −0.050 | 0.025 | −0.099, −0.001 | 0.046 |
| **Maternal PGS-NT × sex (male)** | | −0.090 | 0.036 | −0.160, −0.020 | 0.013 |
| **Paternal PGS-NT × sex (male)** | | 0.013 | 0.035 | −0.056, 0.082 | 0.703 |

**Table S4.** Assortative mating sensitivity analysis: maternal and paternal PGS-NT in the same model, trio subsample (N = 7,302 observations). Selected coefficients shown; full model includes age, sex, birth year, and genotyping array as covariates. PGS-T = transmitted polygenic score; PGS-NT = non-transmitted polygenic score. β = standardised regression coefficient. Sex reference category is female. Maternal PGS-NT main effect represents the effect in females (reference category).
